# Evidence of transmission from fully vaccinated individuals in a large outbreak of the SARS-CoV-2 Delta variant in Provincetown, Massachusetts

**DOI:** 10.1101/2021.10.20.21265137

**Authors:** Katherine J. Siddle, Lydia A. Krasilnikova, Gage K. Moreno, Stephen F. Schaffner, Johanna Vostok, Nicholas A. Fitzgerald, Jacob E. Lemieux, Nikolaos Barkas, Christine Loreth, Ivan Specht, Christopher H. Tomkins-Tinch, Jillian Silbert, Beau Schaeffer, Bradford P. Taylor, Bryn Loftness, Hillary Johnson, Petra L. Schubert, Hanna M. Shephard, Matthew Doucette, Timelia Fink, Andrew S. Lang, Stephanie Baez, John Beauchamp, Scott Hennigan, Erika Buzby, Stephanie Ash, Jessica Brown, Selina Clancy, Seana Cofsky, Luc Gagne, Joshua Hall, Rachel Harrington, Gabrielle L. Gionet, Katherine C. DeRuff, Megan E. Vodzak, Gordon C. Adams, Sabrina T. Dobbins, Sarah D. Slack, Steven K. Reilly, Lisa M. Anderson, Michelle C. Cipicchio, Matthew T. DeFelice, Jonna L. Grimsby, Scott E. Anderson, Brendan S. Blumenstiel, James C. Meldrim, Heather M. Rooke, Gina Vicente, Natasha L. Smith, Katelyn S. Messer, Faye L. Reagan, Zoe M. Mandese, Matthew D. Lee, Marianne C. Ray, Marissa E. Fisher, Maesha A. Ulcena, Corey M. Nolet, Sean E. English, Katie L. Larkin, Kyle Vernest, Sushma Chaluvadi, Deirdre Arvidson, Maurice Melchiono, Theresa Covell, Vaira Harik, Taylor Brock-Fisher, Molly Dunn, Amanda Kearns, William P. Hanage, Clare Bernard, Anthony Philippakis, Niall J Lennon, Stacey B. Gabriel, Glen R. Gallagher, Sandra Smole, Lawrence C. Madoff, Catherine M. Brown, Daniel J. Park, Bronwyn L. MacInnis, Pardis C. Sabeti

## Abstract

Multiple summer events, including large indoor gatherings, in Provincetown, Massachusetts (MA), in July 2021 contributed to an outbreak of over one thousand COVID-19 cases among residents and visitors. Most cases were fully vaccinated, many of whom were also symptomatic, prompting a comprehensive public health response, motivating changes to national masking recommendations, and raising questions about infection and transmission among vaccinated individuals. To characterize the outbreak and the viral population underlying it, we combined genomic and epidemiological data from 467 individuals, including 40% of known outbreak-associated cases. The Delta variant accounted for 99% of sequenced outbreak-associated cases. Phylogenetic analysis suggests over 40 sources of Delta in the dataset, with one responsible for a single cluster containing 83% of outbreak-associated genomes. This cluster was likely not the result of extensive spread at a single site, but rather transmission from a common source across multiple settings over a short time. Genomic and epidemiological data combined provide strong support for 25 transmission events from, including many between, fully vaccinated individuals; genomic data alone provides evidence for an additional 64. Together, genomic epidemiology provides a high-resolution picture of the Provincetown outbreak, revealing multiple cases of transmission of Delta from fully vaccinated individuals. However, despite its magnitude, the outbreak was restricted in its onward impact in MA and the US, likely due to high vaccination rates and a robust public health response.

## INTRODUCTION

On July 10, 2021, the Massachusetts Department of Public Health (MADPH) received reports of an increase in COVID-19 cases among people who reside in or had recently visited Provincetown, MA, a tourist town on Cape Cod. Provincetown had attracted thousands of visitors, many of whom reported attending multiple large public and indoor gatherings, starting with the 4th of July weekend and throughout the following week. COVID-19 incidence in the area rose quickly during the subsequent two-week period, from 0 cases in the 14 days before July 3rd to a peak of 456 cases/100,000 persons/day during July 11th-24th^1^.

Notably, 74% of reported cases were fully vaccinated individuals, of whom 79% were symptomatic ^1^. As of July 1st, estimated COVID-19 full vaccination coverage among the eligible population was 68% in Barnstable County (where Provincetown is located) and 67% across MA^2^. This was the first large, well-characterized outbreak of COVID-19 in a highly vaccinated population in the US^1^, and contributed to the CDC’s decision to reinstate their indoor mask recommendation, including for vaccinated individuals^3^.

The rapid increase in cases despite high vaccination rates prompted state and local public health departments to launch a comprehensive outbreak investigation, including SARS-CoV-2 genome sequencing to characterize the viruses driving the outbreak and to add resolution to contact tracing data. Here we describe the genomic epidemiology of this outbreak, including evidence for SARS-CoV-2 transmission from and between fully vaccinated individuals.

## RESULTS

### SARS-CoV-2 outbreak in a highly vaccinated population

We sequenced SARS-CoV-2 genomes from residual diagnostic specimens collected between July 9th and August 2nd as part of outbreak-associated and enhanced community surveillance testing in the Provincetown area (see Methods for details). We produced high-quality SARS-CoV-2 genomes (unambiguous length ≥24,000 nt and successful gene annotation) from 467 unique individuals. Of these, 439 were epidemiologically confirmed to be associated with the outbreak, representing 40% of the 1,098 known outbreak cases ^4^ (Figure 1A). Genomes were well distributed over the time period of the outbreak (Figure 1B) and closely mirrored the epidemic curve ^1,4^, with densest coverage early when the outbreak was still growing (R_t_>1) (Figure 1C).

**Figure 1.**
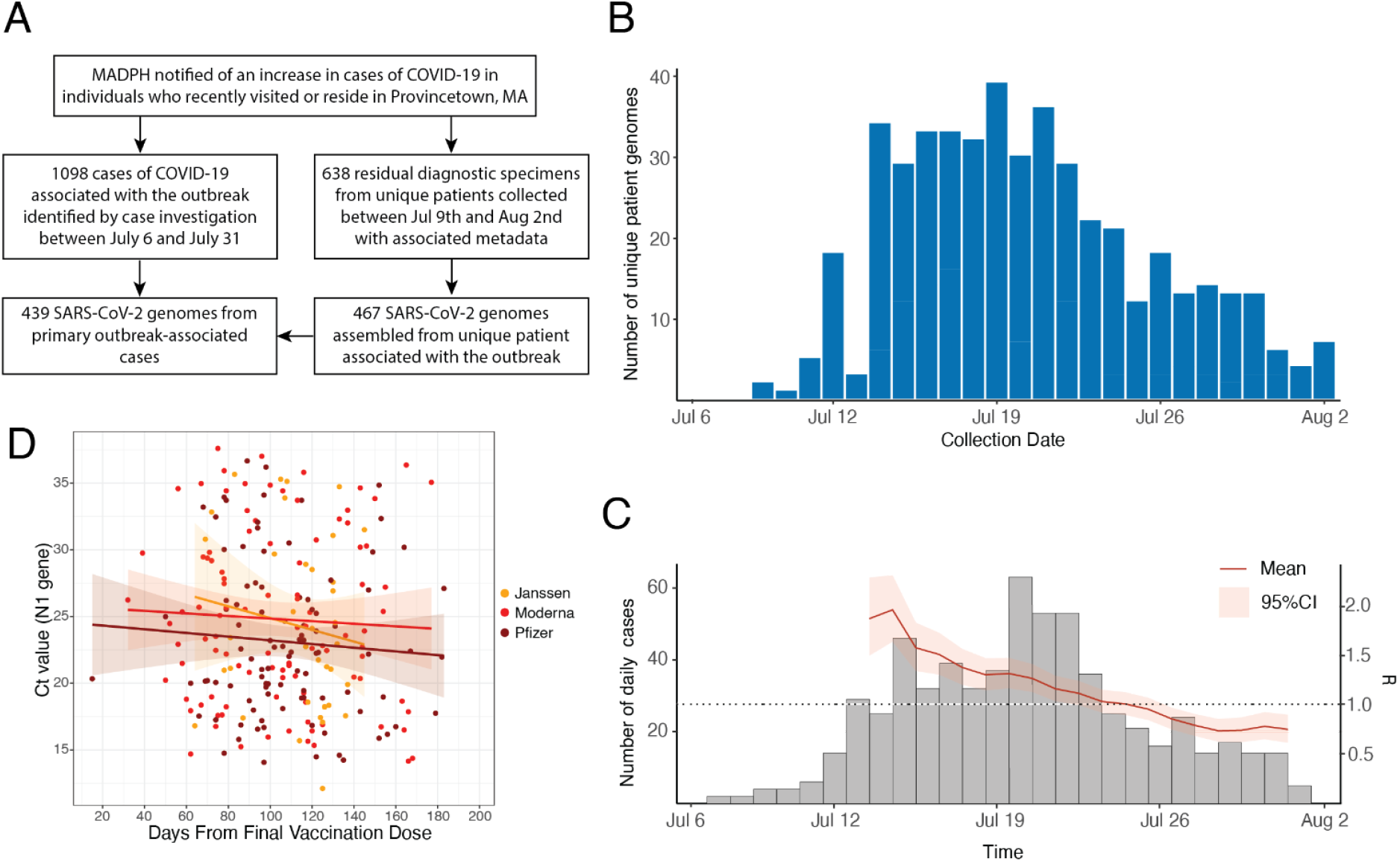
Epidemiology of the Provincetown outbreak and dataset overview. A. Flow diagram describing the main events of the Provincetown outbreak and relevant sample sets presented here. B. SARS-CoV-2 genomes by collection date. Where multiple samples were collected from the same patient, the earliest collection date is used. C. Distribution of all cases in Barnstable county over the time period of the study and estimate of *R*_*t*_ over the course of the outbreak. D. Ct values of the N1 gene for the 313 individuals known to be fully vaccinated by one of Janssen, Moderna, or Pfizer; includes linear regression with 95% confidence interval.

Most specimens obtained for sequencing were from males (80%), the median age was 43, and 84% were from vaccinated individuals, broadly consistent with the outbreak as a whole ^1,4^. Among vaccinated individuals, 48% (203) received Pfizer-BioNTech, 37% (155) Moderna, and 14% (58) Janssen vaccine products. Diagnostic Ct values, an approximation of viral load, were similar between vaccinated and unvaccinated individuals and between symptomatic and asymptomatic individuals, although there were few of the latter because testing focused on symptomatic cases (Figure S1). The average time since completion of the vaccination course was 111 days. Ct values decreased slightly with increasing time since vaccination; however, this trend was not statistically significant in this dataset (Figure 1D). Ct values also decreased slightly with increasing age in vaccinated individuals, though also not significantly. Unvaccinated individuals were on average younger than vaccinated individuals in this cohort (Figure S2).

### Genomic analysis of SARS-CoV-2 viruses from the outbreak

The Provincetown outbreak was driven by the Delta lineage: 99% (462/467) of genomes were Delta, with the remainder being Gamma (Figure 2A). Of the Delta genomes, 84% (394/467) were from the lineage AY.25, a lineage circulating throughout the United States but rarely observed outside North America. The outbreak occurred during a broader rise of the Delta lineage in MA: first detected on March 28th, its prevalence increased sharply in the weeks preceding the July 4th weekend, from 16% on June 15th to 77% at the start of our sampling period on July 9th, and reached 97% by the end of our sampling period on August 3rd (Figure S3).

**Figure 2.**
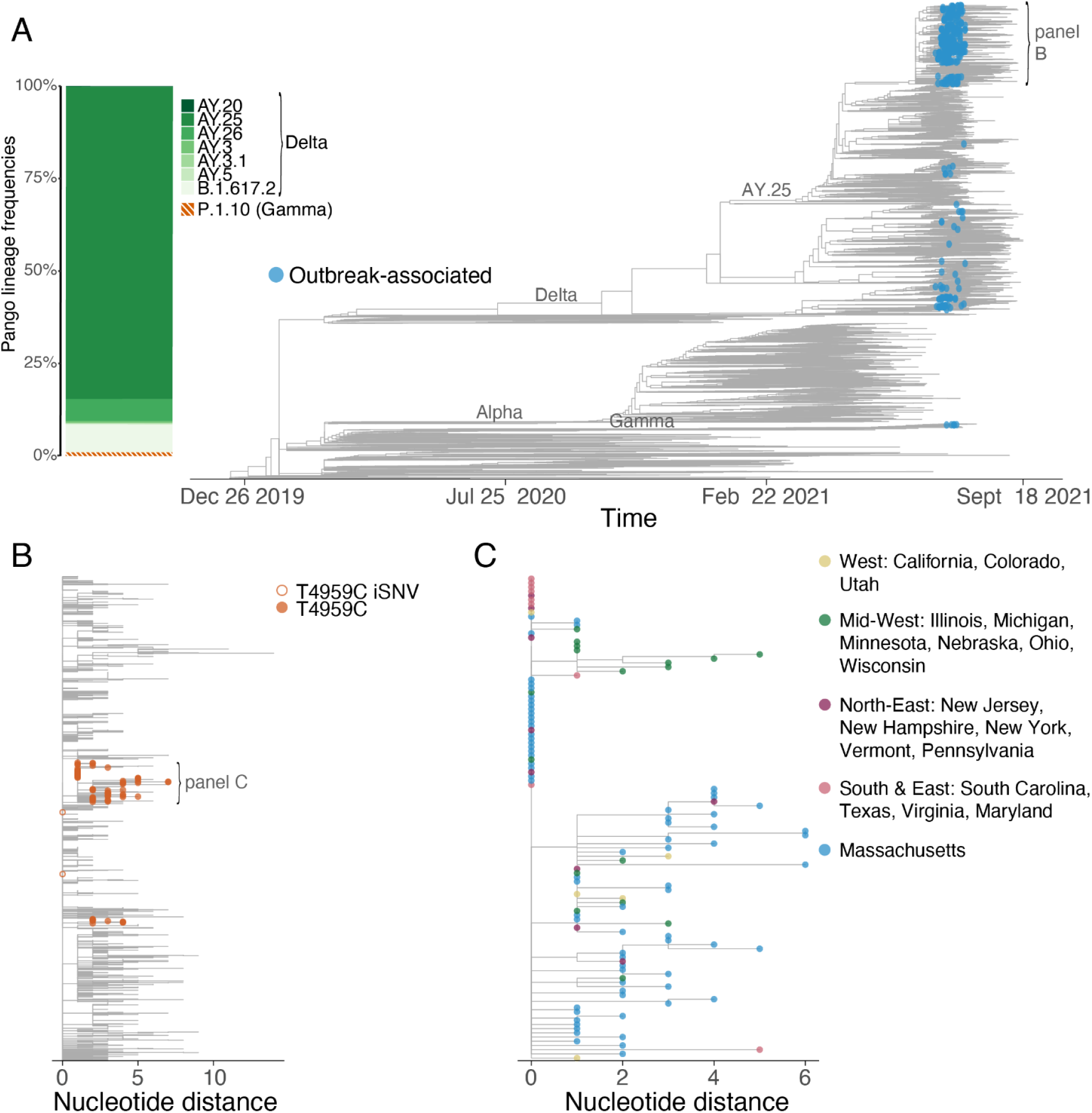
Genomic sequencing of outbreak-associated samples. A. Time tree of SARS-CoV-2 sequences in a global context. The largest cluster of sequences is highlighted by a grey bracket. Inset, frequencies of pango lineages among outbreak-associated genomes. B. A subset of the maximum likelihood tree focused on the largest cluster; samples with the T4959C mutation as either a minor variant (<50%) or consensus variant (50-100%) are highlighted. C. Maximum likelihood tree of the T4959C sub-clade with tips colored by US state, grouped by geographic region.

We found no evidence of genetic differences of known functional consequence between outbreak-associated and other publicly available Delta genomes, or between outbreak-associated genomes from vaccinated and unvaccinated individuals. The Delta genomes in this dataset did not have novel consensus-level variants in the spike protein, nor did they have an increased frequency of any amino acid change of known or suspected functional impact (Figure S4) ^5,6^. Vaccinated individuals also did not have significantly different numbers of intrahost single-nucleotide variants (iSNVs) compared to unvaccinated individuals (p=0.721), suggesting little difference in viral genetic diversity with vaccination (Figure S5A and S5B).

### Phylogenetic analysis of the outbreak

Based on an inferred phylogenetic tree of outbreak-associated SARS-CoV-2 genomes and other publicly available data (Methods), we identified ≥40 distinct branches of the tree in our dataset that predate the outbreak, suggesting that the Delta variant was introduced into this population from many sources. Six of these branches led to clusters (3 or more cases) of varying sizes, of which one cluster—that comprised 83% (387/467) of outbreak-associated genomes—dominated this outbreak (Figure 2A). The remaining 5 clusters each accounted for <4% of primary outbreak-associated cases. The other branches were associated with single cases, each likely representing an independent source of SARS-CoV-2 (Figure 2A and S6). The dominant cluster is defined by 3 nucleotide mutations, C8752T, C20451T, and A26759G, all with no known functional significance. The dominant cluster had an estimated time of most recent common ancestor (tMRCA) of June 18th, 2021 [June 12th - June 24th, 2021]. The earliest reported genomes within the cluster are from other US states, suggesting that this lineage likely emerged outside MA and was introduced into MA just prior to amplification by the outbreak.

A striking feature of the dominant cluster was 158 identical consensus genomes (41% of outbreak-associated genomes in the cluster) at the root of the cluster. This pattern—many identical viral genomes within a short time—usually indicates rapid spread from a single individual and is a signature of superspreading. Public health investigation of the outbreak, however, revealed no evidence for a single exposure site widely shared among cases. Instead, the genomic and epidemiological data taken together suggest that superspreading of the same viral sequence occurred at multiple locations. This is consistent with several scenarios, including one individual infecting others at multiple locations, or several individuals with the same virus, from either a common source or serial infection, transmitting independently. While the specific transmission pattern cannot be resolved, the shape of the phylogeny suggests that overdispersion, in which a few individuals are responsible for most transmission events^7^, continues to play an important role in the COVID-19 pandemic in a landscape dominated by the more transmissible Delta variant^8^.

### Limited spread beyond Provincetown

We investigated the extent to which cases descending from the outbreak contributed to the subsequent increase in cases, largely due to Delta, in MA and across the US. Approximately half of outbreak-associated individuals reported residency in MA, with the remainder visiting from 20 other US states^4^, raising the possibility that the outbreak led to widely dispersed secondary transmission. We first looked for the 3 mutation genomic signature of the dominant outbreak cluster in viral genomes collected after the outbreak as part of ongoing state-wide SARS-CoV-2 genomic surveillance in MA (Methods). We found that the signature of the dominant cluster accounted for a modest and decreasing fraction of Delta infections sequenced in MA over the following weeks, peaking at 9% of all Delta genomes on July 16th and declining to 4% by August 30th (Figure S7). Similar analyses showed that the smaller outbreak clusters had negligible impact on onward spread within the state (not shown).

To quantify the impact of the outbreak more widely in the US, we estimated an upper and lower bound of the spread of the dominant cluster by searching for its descendants in national surveillance data. We estimated an upper bound by identifying all non-MA genomes descended from branches of the dominant cluster with an inferred ancestral origin in MA. Since this set could include viruses from these lineages circulating independently of the Provincetown outbreak, we also estimated a lower bound based on one sub-lineage of the dominant cluster that likely emerged during the outbreak. This sub-lineage is defined by a mutation (T4959C) that first appeared as an iSNV in two individuals from the outbreak (at 39% and 41% frequency), has a tMRCA of July 2nd, 2021 [June 24th - July 7th, 2021], near the start of the outbreak, and was not detected outside of MA until July 13th; we therefore assume all members of the sub-lineage derive from the outbreak (Figure 2B). Based on these two approaches, we infer that the outbreak led to between 44 and 328 sequenced cases in surveillance data from between 18 and 37 states, collected during the period July 10th to September 13th, 2021 (Figure 2C). Among the 37 states, New York, California and Georgia contained the largest number of cases, each representing approximately 10% of the 328 sequenced cases; the distribution of the T4959C mutation was more even. The upper bound represents less than 0.08% of the >400,000 Delta genomes collected in the same period. These findings suggest that, while the outbreak led to some onward transmission, it made at most a modest contribution to later Delta cases within MA and a minimal contribution to cases elsewhere in the US.

### High-confidence transmission from vaccinated individuals

We next investigated the contribution of vaccinated individuals to transmission in the outbreak, using both public health contact tracing and genomic data. Viral genomic data can add resolution to contact tracing by providing an orthogonal measure of the connectivity between cases based on the genetic distance between viruses from infected individuals ^9^. While conventional contact tracing had limited ability to identify direct transmission events in this outbreak, which spanned multiple locations with many possible contacts at each over several days, extensive efforts did identify 19 transmission links with high confidence. These 19 links originated from 17 index cases, 16 of whom were vaccinated^4^. One additional outbreak-associated, vaccinated case was the possible index of a cluster of 19 other cases in a close-contact, residential setting in which all individuals were fully vaccinated. Genomic data was available from both the index and recipient cases for 22 of the 38 putative links identified by contact tracing and, based on the phylogeny, was consistent in every instance with the transmission events inferred by contact tracing, with strong statistical support for 19 (Figure 3A & Figure S8). The phylogeny also supported that all 19 cases in the close-contact cluster stemmed, probably via direct transmission, from a single introduction, possibly from the index case suggested by contact tracing (Figure S9), underscoring the potential for extensive transmission between vaccinated individuals in prolonged close-contact settings.

**Figure 3.**
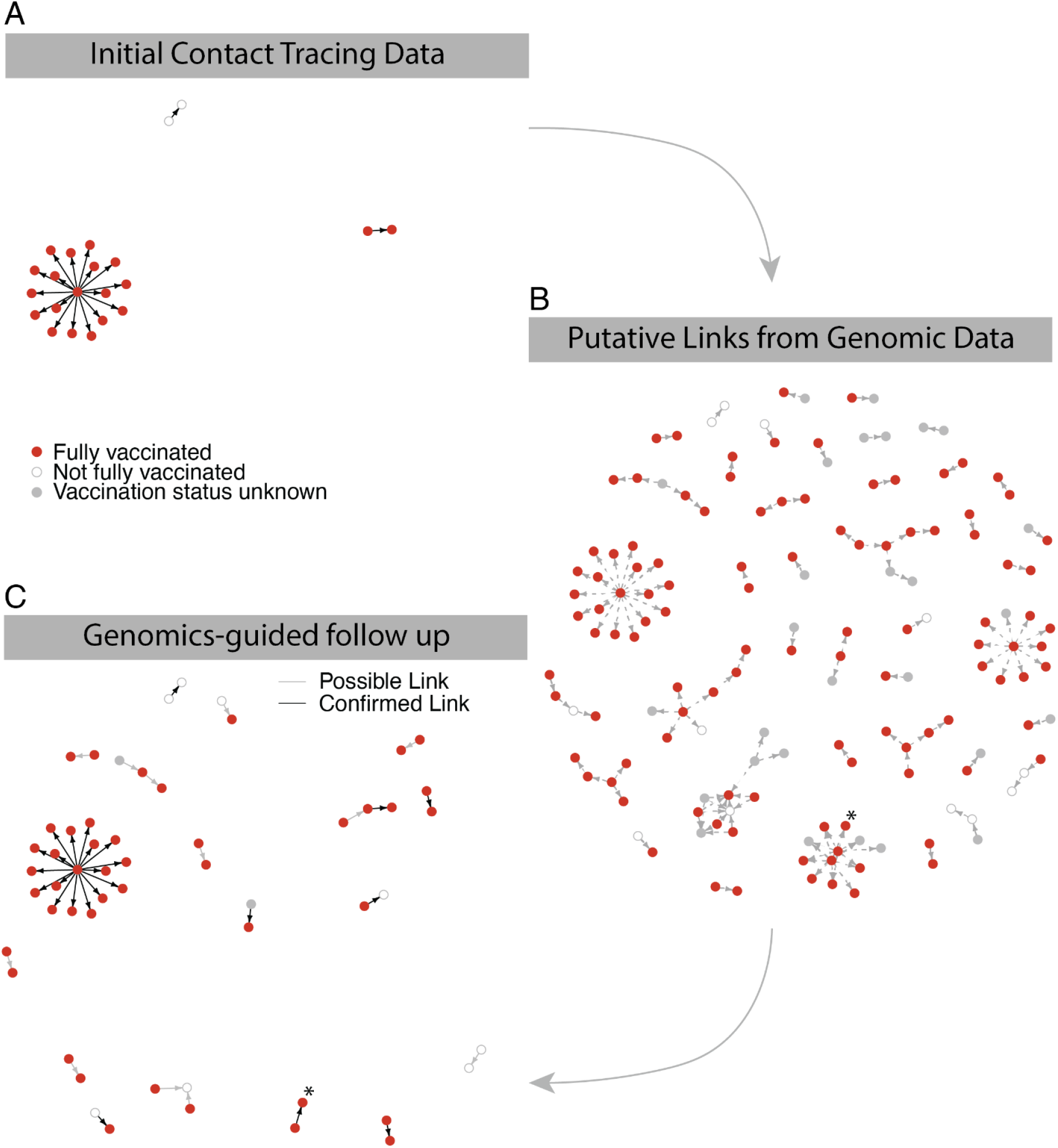
Identification of putative transmission events from vaccinated individuals. A. High-confidence transmission links from contact tracing investigation. B. Predicted transmission links based on genomic sequence, intrahost variants, and symptom onset date. C. Genomics-predicted transmission links corroborated by further epidemiological follow-up. Putative contacts are grouped into possible links (grey lines) and known links (black lines) to denote plausible vs. confirmed contacts between transmission pairs. A confirmed transmission pair that is part of a larger cluster of putative links, described in the text, is marked with an asterisk.

Given the limitations of conventional contact tracing in this outbreak, we applied two empirical methods to infer transmission links based on genomic data (Methods). Using only consensus genomes and temporal information to reconstruct a transmission tree, we identified an additional 56 well-supported links, 38 involving vaccinated index cases (Figure 3B). However, consensus-level genomic data was also limited in its ability to infer transmission links because the low viral diversity in the outbreak often made it difficult to identify direct connections within a cluster. To address this, we used sub-consensus genome diversity (iSNVs) to identify closely related pairs within clusters. This approach can indicate the likely direction of transmission, since variants typically start as iSNVs and become consensus alleles in secondary cases^10,11^. Using iSNVs, we identified an additional 51 putative transmission links, 44 with a vaccinated index case (Figure 3B).

Further epidemiological investigation provided information about 18 of the transmission links identified from genomic data (Figure 3C). In all cases, the additional epidemiological data supported the genomic inference; for the remaining links, contact tracing could neither confirm nor refute the inferred link. Six of the links were classified as involving known contacts and 12 as involving plausible contacts (e.g., living in the same building). In 4 of the 6 known contacts and 9 of the 12 plausible contacts, the putative index case was vaccinated. Most of these predicted links were transmission pairs; however, one putative index case was linked to 9 transmissions based on genomic data (6 to vaccinated individuals). One of these transmissions was identified as being to a known contact, the index having reportedly visited the restaurant where the contact was employed (Figure 3C), raising the possibility of more extensive but unknown transmission from this individual. All of these contacts had been identified by initial contact tracing efforts but were not classified as high confidence transmissions due to the stringency of the epidemiological definition of a link.

Due to the small number of unvaccinated cases in our dataset, we were unable to meaningfully compare rates of secondary transmission by vaccination status (Figure S10). We did note that among vaccinated individuals there were no identified asymptomatic index cases, suggesting a lower risk of transmission from such individuals, but the large uncertainty on the relative risk (95% CI: 0-99% risk of transmission from asymptomatic vaccinated vs. symptomatic vaccinated individual) and possible biases in this observational dataset make drawing conclusions difficult. However, despite the complexities of the epidemiology, contact tracing, and low viral genetic diversity in this outbreak, genomic and epidemiological data combined provide strong support for 21 transmissions from vaccinated individuals and suggestive evidence of a further 9, while genomic data alone provides suggestive evidence for an additional 64. These data suggest that, despite high antibody responses observed for vaccinated individuals from this outbreak^12^, transmission from vaccine breakthrough infections, to both vaccinated and unvaccinated individuals, was common.

## DISCUSSION

The outbreak of SARS-CoV-2 in Provincetown during and after the July 4th weekend was the first large outbreak of the Delta variant in a highly vaccinated population in the US. The robust public health response permitted extensive epidemiological and genomic characterization of the outbreak, the structure of transmission within it, and the role of vaccinated individuals, and offers generalizable insights for containing future outbreaks of Delta and other highly transmissible lineages of SARS-CoV-2.

The Provincetown outbreak raised public health concern and attracted international attention primarily due to the prevalence of symptomatic breakthrough infections and the potential occurrence of transmission from vaccinated individuals. Consistent with other recent reports^13,14^, breakthrough infections with Delta, while often symptomatic and with moderate to high viral loads, were typically mild. Confidently assigning transmission links between individuals was unusually challenging: conventional contact tracing was difficult because of the many locations with dense potential contacts involved, while genomic inference of transmission was hindered by the low overall genetic diversity and large fraction of identical genomes. Nonetheless, using genomic data to prioritize plausible connections between samples followed by more detailed epidemiological investigation identified several likely instances of transmission between fully vaccinated individuals and may serve as a model for future investigations of large outbreaks.

The size of the Provincetown outbreak—over one thousand cases—and its rapid early growth demonstrate that in densely crowded events and indoor conditions the SARS-CoV-2 Delta variant can cause a large outbreak even in a mostly vaccinated population. However, the Provincetown outbreak did not contribute substantially to the increase in Delta cases in Massachusetts. The high rates of vaccination and the swift public health response^1^, which included deployment of mobile testing, a local indoor masking mandate, and an extensive outreach campaign, likely contributed to the short duration and restricted impact of the outbreak. Additionally, the active engagement of the affected community in the epidemiological response, possibly influenced by historical public health outreach in the gay community, may have helped mitigate the impact of the outbreak^15^. The rapid decline and limited impact of the outbreak suggest that while Delta-driven outbreaks are not eliminated by high vaccination rates, they can be controlled with well-understood public health measures.

## Supporting information

GISAID author acknowledgements

## Data Availability

We deposited genomes (Genbank), metadata (BioSample), and raw reads (SRA) to NCBI under BioProject PRJNA715749. All genomes produced in the present study are also available on GISAID.

## Acknowledgements

We thank Patricia Kludt and Meagan Burns from the Massachusetts Department of Public Health; Radhika Gharpure, Samira Sami, Rebecca T. Sabo, Noemi Hall, Anne Foreman, and Scott Laney from the CDC COVID-19 Response Team; the Barnstable County Department of Health and the Environment; the Community Tracing Collaborative; the Boston Public Health Commission; and all members of the local, regional and national COVID-19 emergency response efforts. We also acknowledge all members of the Massachusetts Department of Public Health and Broad Institute SARS-CoV-2 genomic sequencing and surveillance efforts. We gratefully acknowledge the authors from the originating laboratories responsible for obtaining the specimens and the submitting laboratories where genetic sequence data were generated and shared via the GISAID Initiative, on which this research is based (Table S1).

## Funding

This work was sponsored by the Centers for Disease Control and Prevention COVID-19 baseline genomic surveillance contract sequencing (75D30121C10501 to Clinical Research Sequencing Platform, LLC), a CDC Broad Agency Announcement (75D30120C09605 to B.L.M), National Institute of Allergy and Infectious Diseases (U19AI110818 and U01 AI151812 to P.C.S.), the Rockefeller Foundation (2021HTH013 to B.L.M. and P.C.S.), and the Bill and Melinda Gates Foundation (INV-002717 to B.L.M.), as well as support from the Doris Duke Charitable Foundation (J.E.L.), the Howard Hughes Medical Institute (P.C.S.), and the National Human Genome Research Institute (K99HG010669 to S.K.R.). The content is solely the responsibility of the authors and does not necessarily represent the official views of the National Institutes of Health.

## Competing interests

P.C.S. is a co-founder of, shareholder in, and scientific advisor to Sherlock Biosciences, Inc., as well as a Board member of and shareholder in Danaher Corporation. J.E.L. has received consulting fees from Sherlock Biosciences. A.P. is a Venture Partner at Google Ventures. Other authors report no competing interests.

## METHODS

### Sample and data collection

The research project (Protocol #1603078) was reviewed and approved by the MA Department of Public Health Institutional Review Board and covered by a reliance agreement at the Broad Institute. Residual samples were collected and sequenced under a waiver of consent by the MADPH IRB. An additional non-human subjects research and an exempt determination (EX-7080) were made by the Harvard Longwood Campus Institutional Review Board and the Broad Institute Office of Research Subject Protections, respectively, for the analysis of de-identified aggregate and publicly available data. Some study staff maintain dual affiliations with Mass General Brigham and the Broad Institute, but this research was conducted solely at the Broad Institute, Harvard University, and MADPH.

### Epidemiological investigation and case definitions

State and local authorities identified cases linked to the Provincetown outbreak using travel history and exposure data from the state COVID-19 surveillance system and follow-up case investigation. A primary outbreak-associated case was defined as receipt of a positive SARS-CoV-2 test result (nucleic acid amplification test or antigen test) ≤14 days after travel to or residence in Provincetown between July 3rd and July 17th, 2021. The majority of specimens in the present dataset were collected through mobile testing deployed in Provincetown by the MADPH following identification of the outbreak^1^. Primary outbreak-associated cases and cases that did not meet the above criteria as primary cases but were collected in Provincetown or had an epidemiologically confirmed link to a primary case are collectively referred to here as “outbreak-associated”. COVID-19 vaccine breakthrough cases were defined on the basis of either i) documentation from the state immunization registry of completion of COVID-19 vaccination as recommended by the CDC Advisory Committee on Immunization Practices ≥14 days before specimen collection or ii) self-reported vaccination dose(s) indicating completion of COVID-19 vaccination ≥14 days before sample collection during follow-up case investigations. Individuals who had received at least one vaccine dose ≥1 day before sample collection but did not meet these criteria were defined as partially vaccinated.

### SARS-CoV-2 detection and sequencing

We identified specimens for viral genome sequencing linked to the Provincetown outbreak, based on the above criteria and which were submitted to either the Massachusetts State Public Health Laboratory or the Broad Institute for testing, following confirmation by diagnostic RT-qPCR test (Supplemental Methods). Briefly, we extracted RNA from anterior nasal swabs, prepared Illumina sequencing libraries using a multiplexed amplicon approach with the ARTIC V3 primer set (https://artic.network/ncov-2019), and sequenced resulting libraries on an Illumina NovaSeq or MiSeq instrument at the Broad Institute Clinical Research Sequencing Platform or the Massachusetts State Public Health Laboratory (see Supplemental Methods for full protocol descriptions).

### SARS-CoV-2 genome assembly and analysis

Following sequencing, we demultiplexed samples, filtered reads for known sequencing contaminants, and assembled SARS-CoV-2 genomes with NC_045512.2 as reference using publicly available workflows (Supplemental Methods). We deposited assembled genomes meeting CDC criteria for submission to public repositories (unambiguous length ≥24,000 nt and successful gene annotation) in Genbank and GISAID^16^. We deposited genomes (Genbank), metadata (BioSample), and raw reads (SRA) to NCBI under BioProject PRJNA715749. We used LoFreq version 2.1.5 to call intrahost single nucleotide variants (iSNVs) with default parameters (minimum read depth ≥10, strand bias <85%, and default iSNV quality scoring)^17^. Variants with frequency ≥3% and in positions with read depth ≥200 were used for downstream analysis. We constructed phylogenetic maximum-likelihood (ML) and time trees^18^ with associated visualizations using a SARS-CoV-2-tailored Augur pipeline^19^ (*sarscov2_nextstrain_aligned_input*), part of the Nextstrain project ^20^, adapted from github.com/nextstrain/ncov, with the entirety of ARTICv3 amplicons 64, 72, and 73 (Delta dropout regions) masked from tree construction. We included contextual data from the Genbank database (downloaded October 1st, 2021) using two subsampling schemes to prioritize genomes genetically, geographically, and temporally close to outbreak-associated genomes (Supplemental Methods).

### Estimation of transmission events

We used the R package outbreaker2 (version 1.1.2)^21^ to estimate the probability of direct transmission between each pair of individuals using consensus genome sequences. We used the sample collection date where symptom onset date was not available. We used the most complete genome assembly and the earliest collection date as input for individuals with multiple samples. We supplied a Gamma distribution based on a previously described mean and standard deviation of the Delta (B.1.617.2)-specific generation interval and incubation period^22^. We left all other input parameters as unknowns to be estimated by outbreaker2, and allowed for transmission events from individuals not included in the dataset. We ran the outbreaker2 MCMC algorithm 6 times with 1,000,000 iterations each, discarding the first 10% as burn-in. We defined well-supported transmission events as those with mean probability >70%. For the iSNV analysis we defined a putative transmission when 1) the detected iSNV frequency in the contact was ≥50%, 2) the index and contact consensus sequences only differed by 1 consensus-level change, 3) the contact appeared downstream of the index in a divergence phylogeny, and 4) the index’s symptom onset date was ≥2 days before that of the contact. In general, outbreaker2 was more likely to identify transmission pairs, while the iSNV approach identified clusters of individuals.

## Notes

### Author Declarations

This work was reviewed and approved by the Massachusetts Department of Public Health Institutional Review Board and covered by a reliance agreement at the Broad Institute.

