## Supplemental Material for "Evidence of transmission from fully vaccinated individuals in a large outbreak of the SARS-CoV-2 Delta variant in Provincetown, Massachusetts"

Supplemental Materials

### Table of Contents


| | |
|---|---|
| Supplemental Methods | 3 |
| Figure S1 | 8 |
| Figure S2 | 9 |
| Figure S3 | 10 |
| Figure S4 | 11 |
| Figure S5 | 12 |
| Figure S6 | 13 |
| Figure S7 | 14 |
| Figure S8 | 15 |
| Figure S9 | 16 |
| Figure S10 | 17 |
| Table S1 | 18 |
| References | 18 |

### Supplemental Methods

**SARS-CoV-2 detection and sequencing methods**

For specimens submitted to the Broad Institute, total RNA was extracted from inactivated AN swabs using the Thermo Fisher MagMAX Viral RNA Isolation kit and presence of virus was confirmed by RT-qPCR assay detecting the N1 and N2 gene regions of the virus under Emergency Use Authorization at the Broad Institute Clinical Research Sequencing Platform in a CLIA-compliant diagnostic laboratory. Ct values for the N1 gene were used to compare viral titers between individuals; samples for which the RP positive control gene had a Ct>32 were excluded from the analyses to prevent biasing towards high viral loads.

Following a positive (N1 and N2 detected) or inconclusive (only one of N1 or N2 detected) test result, candidate samples were re-extracted from the source material and Illumina sequencing libraries were prepared using the NEBNext ARTIC v3 SARS-CoV-2 FS Library Prep Kit. Libraries were sequenced on Novaseq SP flowcells with 75-nucleotide paired-end reads. During library preparation, some volumes were adjusted from manufacturer recommendations to accommodate 384-well plate reactions and high-throughput automated processing.

For specimens submitted to the MADPH, total RNA was extracted using the Roche MagNA Pure Total Nucleic Acid Isolation kit. Following extraction, samples proceeded to cDNA synthesis, amplification using multiplex PCR primers, and preparation with Illumina DNA Prep. The DNA libraries were purified and denatured before hybridization of the biotin probe oligonucleotide pool in preparation for sequencing. Libraries were sequenced on Illumina MiSeq with 2x150 paired-end reads.

**SARS-CoV-2 genome assembly approaches and code availability**

For sequences generated at the Broad Institute we conducted all analyses using viral-ngs 2.1.28 on the Terra platform (app.terra.bio). All of the workflows named below are publicly available via the Dockstore Tool Registry Service (dockstore.org/organizations/BroadInstitute/collections/pgs). Briefly, samples were demultiplexed, reads were filtered for known sequencing contaminants, and SARS-CoV-2 genomes were assembled using a reference-based assembly approach with the reference genome NC_045512.2 (*sarscov2_illumina_full.wdl)*. For sequences generated at the MADPH, all analyses were executed on a local, on-premise, Linux compute machine at the Massachusetts State Public Health Laboratory (MASPHL). We processed all raw read data using a reference-based consensus calling method with the SARS-CoV-2 isolate Wuhan-Hu-1 genome (NC_045512.2) as reference. The workflow is publicly available on GitHub (github.com/AndrewLangvt/genomic_analyses/blob/main/workflows/wf_viral_refbased_assembly.wdl).

Assembled genomes meeting the CDC criteria for submission to public repositories (unambiguous length ≥24,000 nt and successful gene annotation) were deposited in NCBI Genbank and GISAID[1]. Raw reads for all samples (including those that did not produce a successful genome) were deposited in NCBI SRA. All NCBI data were deposited under BioProject PRJNA715749 and have been tagged with the BioSample attribute purpose_of_sequencing, set to a value of "Cluster/Outbreak investigation" for primary and secondary outbreak-associated cases identified by MA DPH epidemiologists or "Targeted surveillance (non-random sampling)" for samples collected as part of enhanced surveillance efforts but where no primary link to the outbreak was known. In the main text, both of these groups are together referred to as outbreak-associated. Where an individual received multiple positive tests, we used for analysis the most complete genome that met all criteria to be publicly shared (if two or more genomes were of the same length, we selected the genome from the earlier collection time). We confirmed that genomes generated from the same patient were concordant. Genome pairs from two individuals differed by a single mutation and a pair of genomes from one individual differed by two mutations. These mutations did not impact phylogenetic assignment or other inferences and likely result from lower coverage in one of the pairs.

##### Phylogenetic Tree subsampling strategies

In order to identify likely sources of introduction or export, we included contextual data from the Genbank database (downloaded August 13th, 2021) using two approaches. First, we used a focal weighted subsampling scheme (nextstrain priorities.py) to prioritize genomes genetically, geographically, and temporally close to our outbreak-associated genomes (https://github.com/broadinstitute/nextstrain-builds/blob/main/builds/broad-usa-builds.yaml#L125). Second, in addition to this weighted subsampling scheme, we forced inclusion of a set of contextual genomes identified by phylogenetic proximity by concatenating: (a) a list of samples obtained by performing a sequence search in UCSC UShER[2] (against the "GISAID, Genbank, COG and CNCB (3,960,091 sequences)" database on September 28th, 2021, with "Number of samples per subtree showing sample placement" set to 100), resulting in the identification of 3,970 proximal samples, and (b) a list of samples obtained by constructing a phylogenetic tree using FastTree[3,4] (version 2.1.11) on a masked multiple sequence alignment (retrieved from GISAID August 20th, 2021) of a random sample of 194,716 Delta lineage viral sequences, using iterative tree refinement followed by a greedy depth-first search to identify outbreak-enriched clades (>10% of total leaf nodes being outbreak samples). Concatenation of the above lists as well as the samples of interest in this study, followed by deduplication, resulted in forced inclusion of 6,372 samples.

##### Ongoing presence of outbreak-associated mutations

We used the contextualized maximum likelihood phylogeny to estimate the number of introductions that seeded the Provincetown outbreak and exports descending from the Provincetown outbreak. Using Nextstrain's[5] ancestral inference, we inferred the association of each internal node to the Provincetown outbreak. We defined an introduction as an ancestral trait change from not Provincetown-associated to Provincetown-associated. Using baltic (https://github.com/evogytis/baltic), we traversed the phylogeny to find changes in state of internal nodes from not Provincetown-associated to Provincetown-associated with an inferred date of June 15th or later. Each of these introductions, with the resulting cluster, were pulled out and plotted using matplotlib. Similarly for exports, we inferred the association of each internal node to the division in which it was collected. We defined exports as changes in inferred geographic division starting from a branch inferred to be from Massachusetts. To find exports from the dominant cluster of the Provincetown outbreak, we started at tips that were associated with the outbreak. We traversed the tree from these tips upward in the hierarchy until the earliest internal node with an inferred date on or after July 3rd; from that collection of nodes we traversed the tree towards later time points and catalogued all nodes that were not located in Massachusetts as an upper bound to downstream exported transmissions.

To quantify the impact of the Provincetown outbreak on subsequent spread of Delta lineage viruses in Massachusetts and identify novel or frequent mutations of functional consequence, we compared the frequency of mutations detected in outbreak-associated genomes to their frequency in publicly available data. We downloaded from GISAID[1] all MA Delta genomes with collection date between July 3rd and August 30th, 2021, and purpose of sequencing listed as “baseline surveillance.” All sequences were processed with Nextclade CLI version 1.3.0 and custom python code. We used nucleotide substitutions that define each of the five clusters within Delta to count the number of publicly available sequences in each cluster and used collection date to calculate a daily frequency for each cluster compared to all baseline surveillance Delta genomes.

**Estimate of effective reproductive number**

We used a simple model using the parametric_si method in the R package EpiEstim v4.0.1[6] to estimate the effective reproductive number ($R_t$), the average number of secondary cases per infectious case at a given time, for the Provincetown outbreak using case counts of all cases in MA associated with this outbreak with specimen collection dates from July 6th through July 31st, 2021. Our estimates assume that Delta has a serial interval of 2.3 days with a standard deviation of 3 days[7]; that the serial interval is the same for vaccinated and unvaccinated individuals; and that there are no negative serial intervals, where a contact becomes symptomatic before the index. The expression used to calculate $R_t$ is $R_t = c(t)/c(t - \tau)$, where $c(t)$ is the incidence at time $t$ and $\tau$ is the mean value of the serial interval.

**Estimate of transmission rates**

To assess differences in transmission rates between vaccinated and unvaccinated individuals, we counted individuals in the two categories who were index cases in well-supported transmissions and used those counts to calculate the relative risk of transmission. A large cluster of secondary cases associated with a known close-contact setting was excluded from the analysis. We estimated the relative risk and calculated confidence intervals by constructing via simulation the likelihood function for the observed number of transmissions from the two categories, based on the number of samples in each category, under a model with one free parameter, the relative risk of transmission. In the model, error in inferring the index case was accommodated by replacing the vaccination status of a fraction of true index cases with the status of a sample drawn randomly from the population; the probability of replacement was itself drawn from the distribution of estimated uncertainties in index case assignment. Four million iterations of the simulation yielded a maximum likelihood estimate of the relative risk and a 95% confidence interval (determined from a decrease in the likelihood of 1.92 logs). A similar procedure was used for symptomatic/asymptomatic transmission from vaccinated individuals, with 100,000 iterations of the simulation.

#### Supplemental Figures and Tables

**Figure S1. Symptoms, Ct, and vaccination status.** Ct values in outbreak-associated cases (465 individuals passing Ct thresholds; see Methods). In individuals with multiple samples, the earliest collected sample was used. Presence or absence of symptoms was known for 263 individuals; of these, vaccination status was known for 251. Partially vaccinated individuals were excluded from the analysis at right. In each distribution, the mean is shown by a red line; the mean ± one standard deviation is shown by dashed red lines.

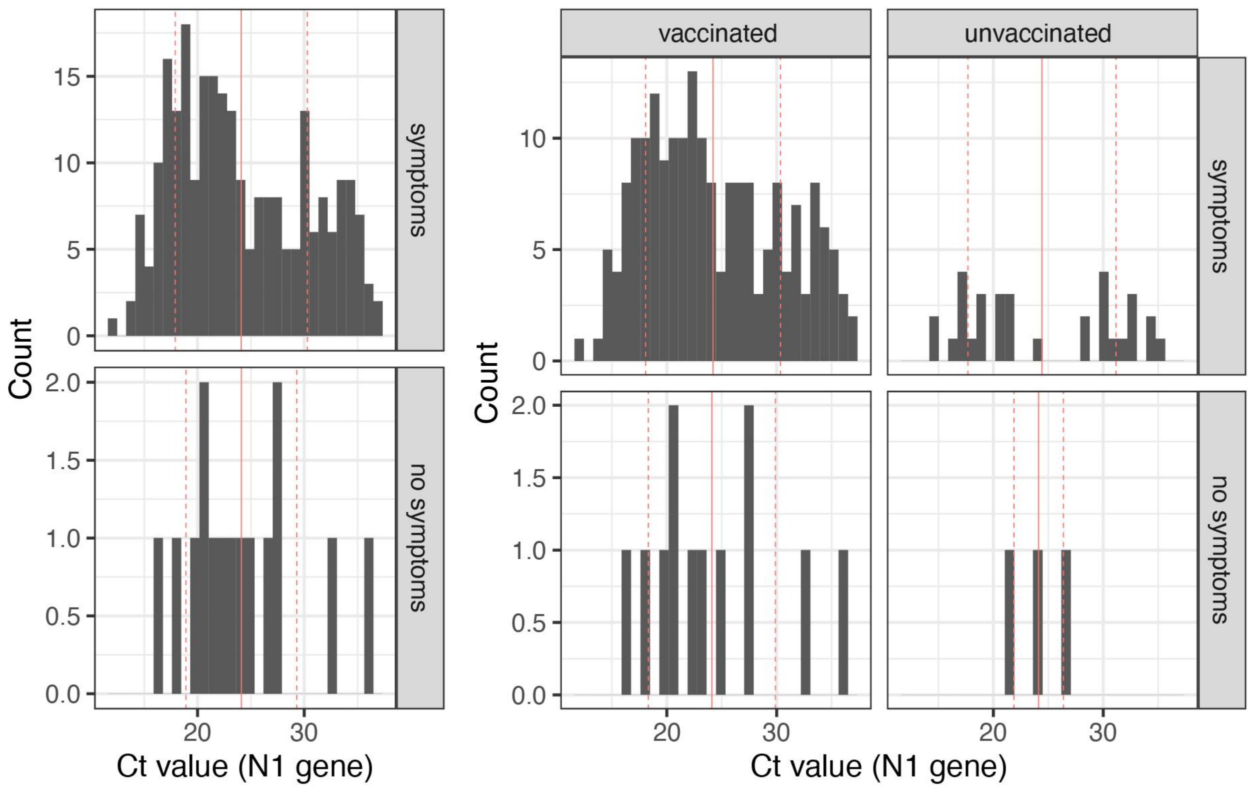

**Figure S2. Age, Ct, and vaccination.** Ct values by age and vaccination status and age distributions by vaccination status and vaccine brand in 465 outbreak-associated cases passing Ct thresholds (see Methods). In individuals with multiple samples, the earliest collected sample was used. Individuals with unknown vaccination status and partially vaccinated individuals are excluded from all three panels. Vaccination status was known for 355 individuals; of these, all had a known age and 290 were known to be fully vaccinated by one of Janssen, Moderna, or Pfizer. In each distribution, the mean is shown by a red line; the mean ± one standard deviation is shown by dashed red lines. Scatterplot includes linear regression with 95% confidence interval.

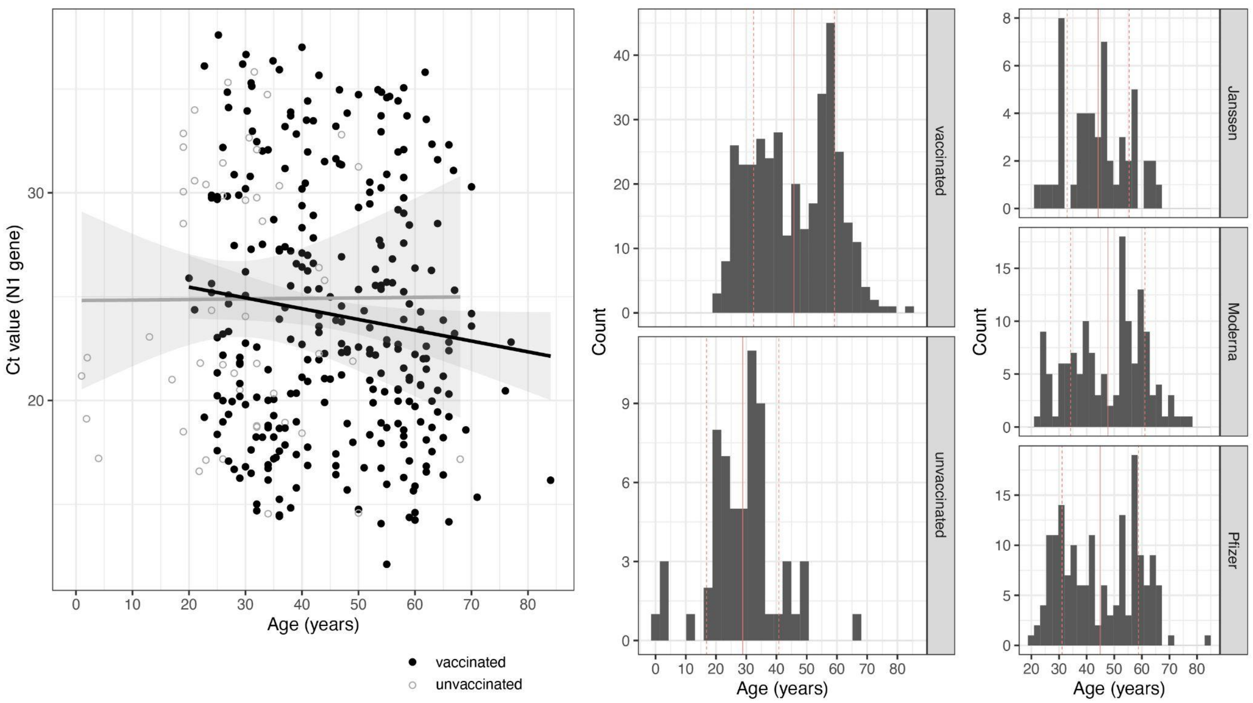

**Figure S3. Frequency of Delta lineages in Massachusetts.** The proportion by epidemiological week of Delta lineage sequences among all publicly available baseline surveillance data from Massachusetts. Data shown is only that generated by the Clinical Research Sequencing Platform and Viral Genomics Group at the Broad Institute.

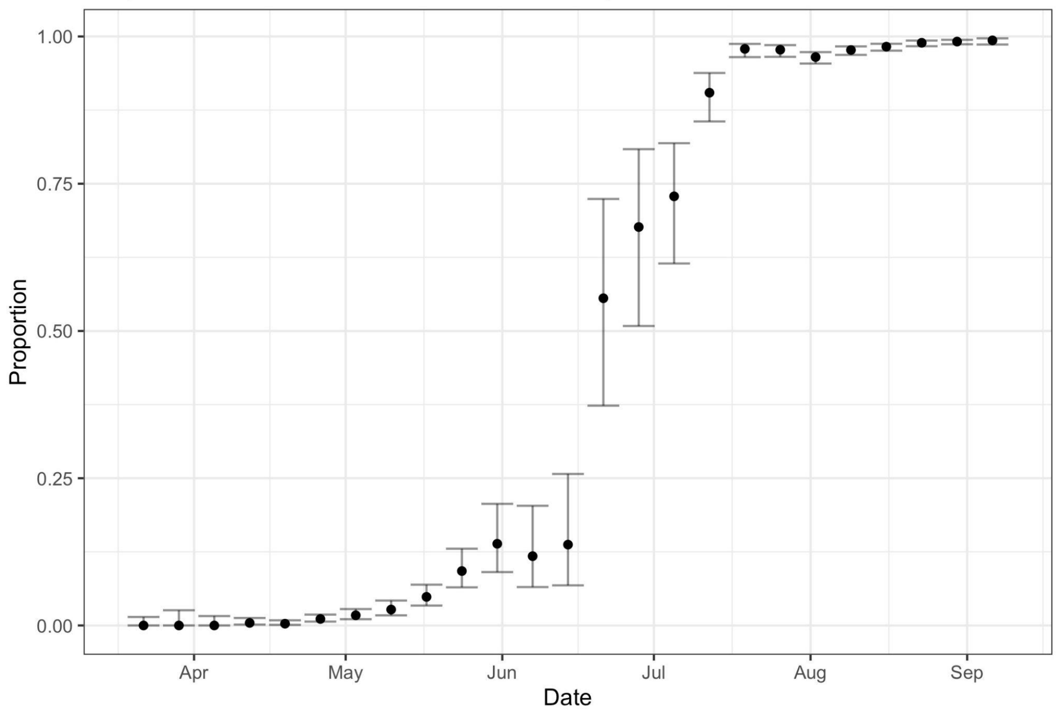

**Figure S4. SARS-CoV-2 consensus variants in outbreak-associated genomes.** The frequency of the 50 most common consensus-level mutations among all outbreak-associated genomes (blue) compared to the proportion of Delta genomes in GISAID with the same mutation (grey). All AY.25 genomes had an amino acid change at position E239Q in ORF3a; however, although rare among publicly available Delta genomes, E239Q is shared across the AY.25 lineage and is of no known functional significance.

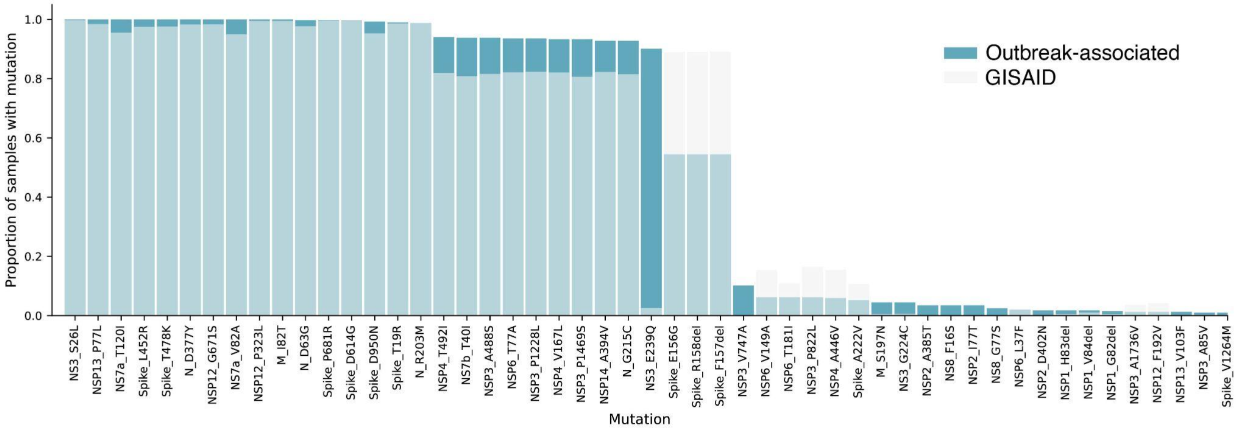

**Figure S5. Intrahost variant frequencies in vaccinated and unvaccinated individuals.** A. Total number of iSNVs per individual grouped by vaccination status. B. The number of observations of each iSNV across all samples. iSNVs are labelled by their gene and amino acid change (if nonsynonymous) or nucleotide position (if synonymous). Bars are colored by the vaccination status of each individual in which a mutation was observed.

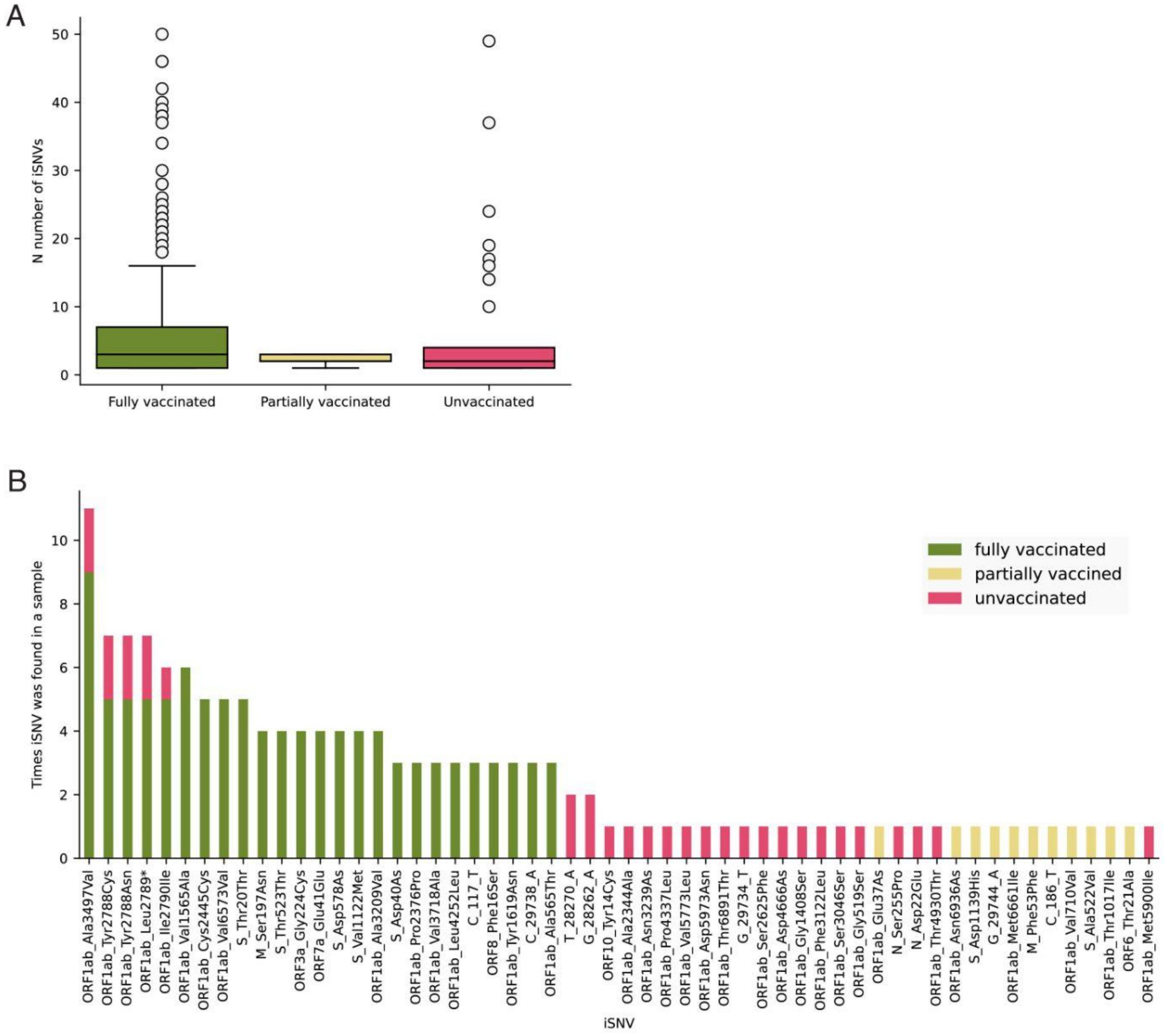

**Figure S6. Estimation of introductions into the outbreak.** A) Time tree of SARS-CoV-2 sequences in a global context (as in Figure 2A), colored by association with the Provincetown outbreak. B) Plot of each introduction into Provincetown as inferred from the phylogenetic tree based on a change in ancestral inference of a node to "outbreak-associated." Large, gray dots represent the estimated tMRCA of the clade from outside of Provincetown. All outbreak-associated samples downstream of each node are shown as in A.

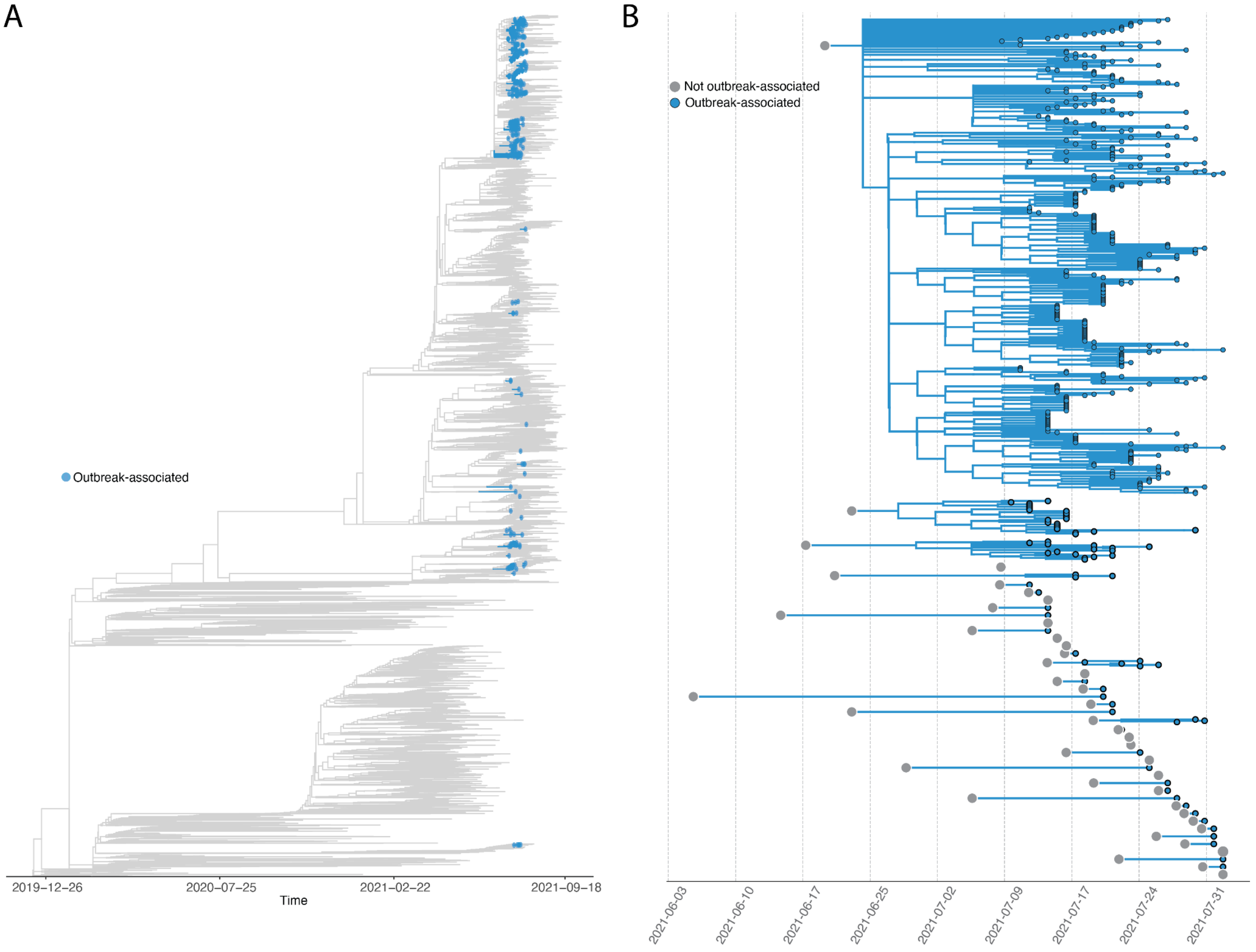

**Figure S7. Frequency of the mutational signature of the largest cluster with evidence of community spread in Massachusetts genomes.** Plot shows the percentage of all Delta lineage baseline surveillance genomes from MA with the mutational signature of the dominant cluster among outbreak-associated cases. Three mutations (C8752T, C20451T, and A26759G) are shared by the majority of all outbreak-associated genomes.  Percentages per day based on sample collection date and new cumulative cases in MA were plotted over time from July 3rd - August 31st, 2021.

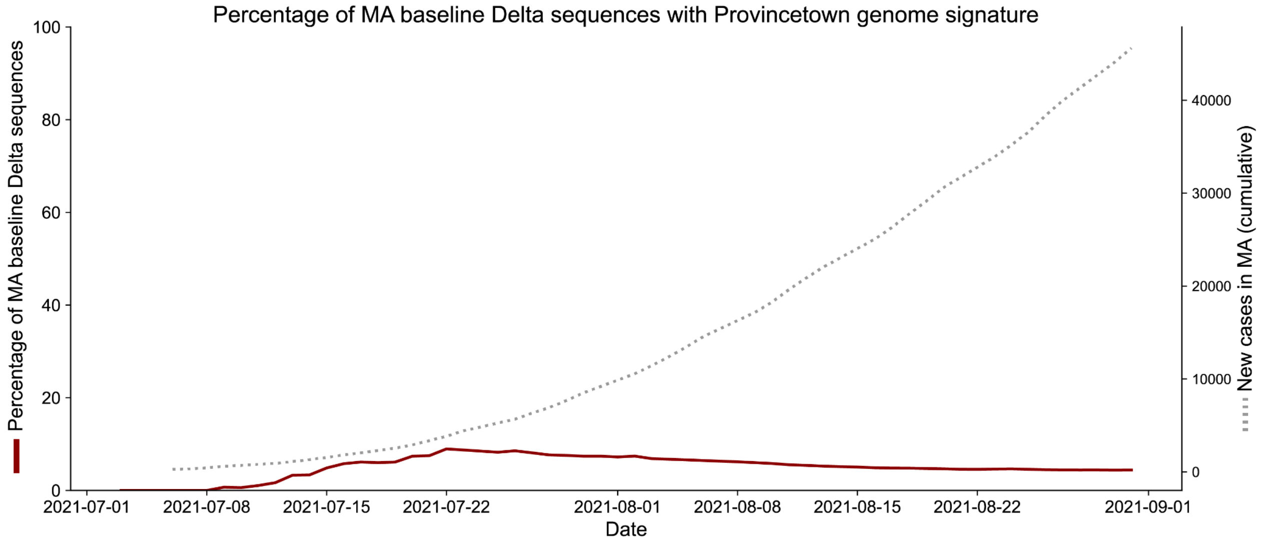

**Figure S8. Limitations of genomic prediction of transmission links.** Maximum likelihood phylogenetic tree of the only high-confidence transmission pair from contact tracing without strong statistical support in outbreaker2 transmission reconstruction. This pair was in a cluster of 6 identical consensus genomes with very similar collection dates. No symptom onset date was known for either of these individuals. Genomic data was thus consistent with transmission but this link was not predicted based on genomic and temporal data alone. Indeed, even when incorporating contact tracing information into the model, another sample was predicted as almost equally likely to have been the ancestor of this case (frequency=43.8% compared to 49.8% for the epidemiologically determined index). This image is part of a larger phylogenetic tree available at [https://auspice.broadinstitute.org/sars-cov-2/ma-delta/20211005/cluster-unique-usher](https://auspice.broadinstitute.org/sars-cov-2/ma-delta/20211005/cluster-unique-usher)

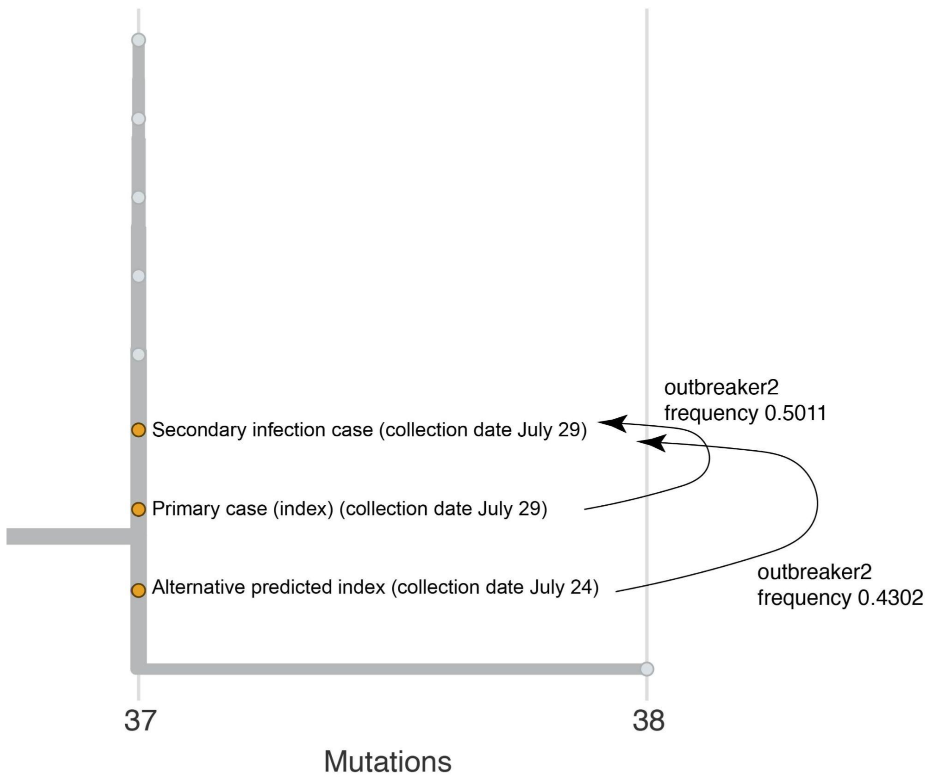

**Figure S9. Phylogenetic placement of known high-confidence transmission links determined through contact tracing.** Maximum likelihood phylogeny of the large cluster of cases associated with a single index case in a close-contact setting. Known contacts colored in dark red. This image is part of a larger phylogenetic tree available at https://auspice.broadinstitute.org/sars-cov-2/ma-delta/20211005/cluster-unique-usher

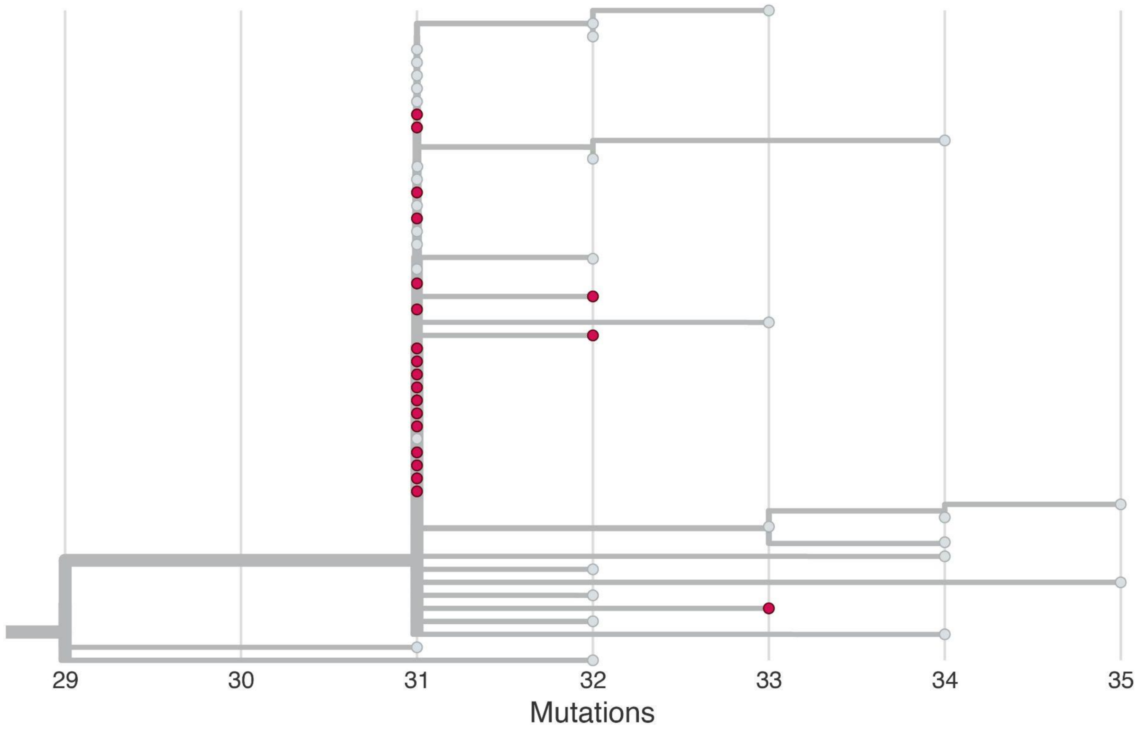

**Figure S10. Transmission by vaccination and symptoms.** Grey bars, fractions, and 90% exact binomial confidence intervals indicate the proportion of individuals that were the origin of at least one transmission event predicted by outbreaker2 with probability >70%, divided by vaccination status (left) and presence or absence of symptoms in vaccinated individuals (right). 90% binomial confidence intervals were calculated using the exact method through the binom package in R. Using simulations incorporating outbreaker2's confidence in putative transmission links (see Supplemental Methods), we calculate that an infected unvaccinated individual was 0.18-2.11 times as likely to transmit as an infected vaccinated individual. Among fully vaccinated individuals, an infected asymptomatic individual was 0-0.99 times as likely to transmit as an infected symptomatic individual. Our estimates of relative risk are predicated on outbreaker2 correctly estimating the probability that it has chosen the right index case of each putative transmission.

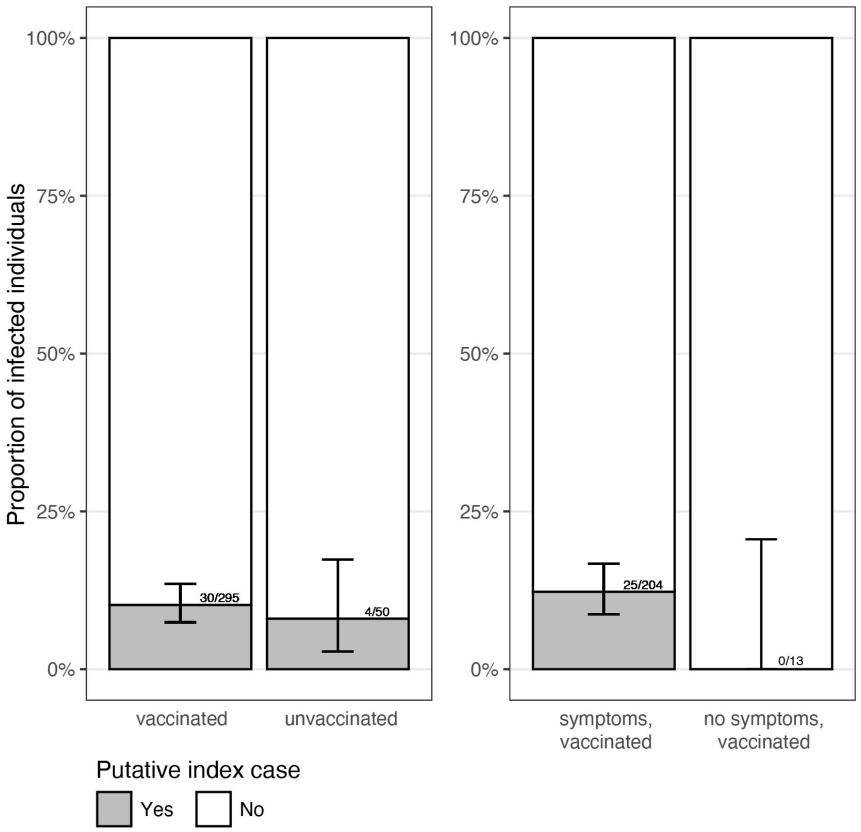

**Table S1.** Acknowledgement table of GISAID data contributors [provided as additional text file]
